# Comparison of Local Influenza Vaccine Effectiveness Using Two Methods

**DOI:** 10.1101/2020.09.14.20194449

**Authors:** GK Balasubramani, Richard K Zimmerman, Heather Eng, Jason Lyons, Lloyd Clarke, Mary Patricia Nowalk

## Abstract

**Background:** In some settings, research methods to determine influenza vaccine effectiveness (VE) may not be appropriate because of cost, time constraints, or other factors. Administrative database analysis of viral testing results and vaccination history may be a viable alternative. This study compared VE estimates from outpatient research and administrative databases.

**Methods:** Using the test-negative, case-control design, data for 2017-2018 and 2018-2019 influenza seasons, were collected using: 1) research methods including consent, specimen collection, RT-PCR testing and vaccine verification using multiple methods; and 2) an administrative database of outpatients with a clinical respiratory viral panel combined with electronic immunization records. Odds ratios for likelihood of influenza infection by vaccination status were calculated using multivariable logistic regression. VE = (1 - OR) × 100.

**Results:** Research participants were significantly younger (*P*<0.001), more often white (69% vs. 59%; *P*<0.001) than non-white and less frequently enrolled through the emergency department (ED) (35% vs. 72%; *P*<0.001) than administrative database participants. VE was significant against all influenza and influenza A in each season and both seasons combined (37%-49%). Point estimate differences between methods were evident, with higher VE in the research database, but insignificant due to low sample sizes. When enrollment sites were separately analyzed, there were significant differences in VE estimates for all influenza (66% research vs. 46% administrative *P*<0.001) and influenza A (67% research vs. 49% administrative; *P*<0.001) in the ED.

The selection of the appropriate method for determining influenza vaccine effectiveness depends on many factors, including sample size, subgroups of interest, etc., suggesting that research estimates may be more generalizable. Other advantages of research databases for VE estimates include lack of clinician-related selection bias for testing and less misclassification of vaccination status. The advantages of the administrative databases are potentially shorter time to VE results and lower cost.

## Introduction

Since 2011, the US FLU VE Network has been estimating influenza vaccine effectiveness (VE) using a test negative design that requires specimen collection from patients seeking medical care for an acute respiratory illness. Recruitment takes place in outpatient settings such as urgent care centers, primary care offices and emergency departments. Consenting, enrolling and swabbing for PCR testing for presence of influenza is frequently performed by research personnel who are not members of the clinical staff.

With the onset of the COVID-19 pandemic, the ability to enroll patients with acute respiratory illness was abruptly curtailed. Many institutions temporarily ceased all but essential research, personal protective equipment (PPE) was in short supply and was being reserved for the protection of health care workers, while vast amounts of resources, human and otherwise, were diverted to the containment, treatment and prevention of SARS-CoV-2 infection.

In addition, the ability to test for influenza and other respiratory viruses locally, was hampered by the health system’s decision to prioritize SARS-CoV-2 testing over respiratory viral panels (RVPs) (Graham Snyder, MD, personal communication, 2020). The timeline for adding SARS-CoV-2 to the currently used multi viral testing platforms, and the duration and severity of the novel coronavirus pandemic remain unknown. It is unlikely that SARS-CoV-2 will displace influenza in the coming influenza season and the viruses will most likely co-circulate. Given that the need to determine influenza vaccine effectiveness continues, other methods of determining influenza vaccine effectiveness should be explored. Previous studies have used administrative databases to estimate influenza outpatient visits,^1^ track influenza outbreaks^2^ and calculate influenza VE in specific population subgroups such as pregnant women^3^ and older adults.^4^

In this study, VE estimates using methodology from the US Flu VE Network (research database) are compared with VE estimates using data from a clinical surveillance software system (administrative database). The advantages and disadvantages of both methods of estimating influenza VE are discussed.

## Methods

### Data

Data used for this analysis were collected from three sources: 1) a local health system’s clinical surveillance software system which extracts virology test results from the EMR (Theradoc); 2) the Pennsylvania Statewide Immunization Information System (PA-SIIS); and 3) research data from local outpatient facilities and emergency departments participating in the US Flu VE Network.

An IRB-approved honest broker extracted data from Theradoc on a cohort of Allegheny County residents who received an outpatient RVP test at a hospital-based clinic or emergency department of one of the general acute care hospitals in the health system during the study period that included the 2017-2018 and 2018-2019 influenza seasons. For patients with more than one visit with RVP tests ≤14 days apart, only data from the first visit were included. If visits occurred greater than 14 days apart, then data from both visits were included.

Analysts created annual Boolean indicators for influenza from the RVP results. If no positive influenza result was observed, the final specimen collection date was saved to confirm there was no infection as of that date. This list also contained basic demographic data of race, sex and age. This list of patients was combined with immunization records from the Pennsylvania State immunization Information System (PA-SIIS). In cases where an individual had more than one vaccination in a given influenza season, the immunization records were reduced to a single record per patient per influenza season by selecting the first vaccination date. This dataset is henceforth called the “administrative” database.

The “research” database was derived from participants who were recruited from ambulatory, urgent care clinics and emergency departments during the 2017-2018 and 2018-2019 influenza seasons for the US Flu VE Network study. Detailed study methods on the US Flu VE Network have been described elsewhere.^5-9^ Briefly, patients aged ≥6 months presenting with an acute respiratory infection (ARI) including cough within 7 days of symptom onset were enrolled at participating outpatient healthcare facilities, including community physician offices, urgent care centers and emergency departments. Patients who had received antiviral medication in the 7 days before enrollment or had been enrolled in the prior 14 days were ineligible. Following informed consent, study staff collected respiratory specimens (nasal and throat swabs from patients aged ≥2 years or nasal swabs only from patients aged <2 years) for influenza virus testing (including virus type and subtype) by reverse-transcription polymerase chain reaction (RT-PCR). Demographic data were obtained from interview. Vaccination status was based on documented receipt of each year’s influenza vaccine from PA-SIIS.

The two databases were considered to be independent because they primarily included patients from different clinical sites and because enrollees who had clinical RVP testing and were enrolled in the US Flu VE network study accounted for <4% of the total administrative database.

### Study Periods

The influenza circulation period, defined as the dates between the first and last influenza positive research enrollment during each season, was determined for each year in both the administrative and research databases. Subjects with influenza testing performed outside the influenza circulation periods were excluded from analyses. The enrollment period details are shown in Table 1.

**Table 1.**
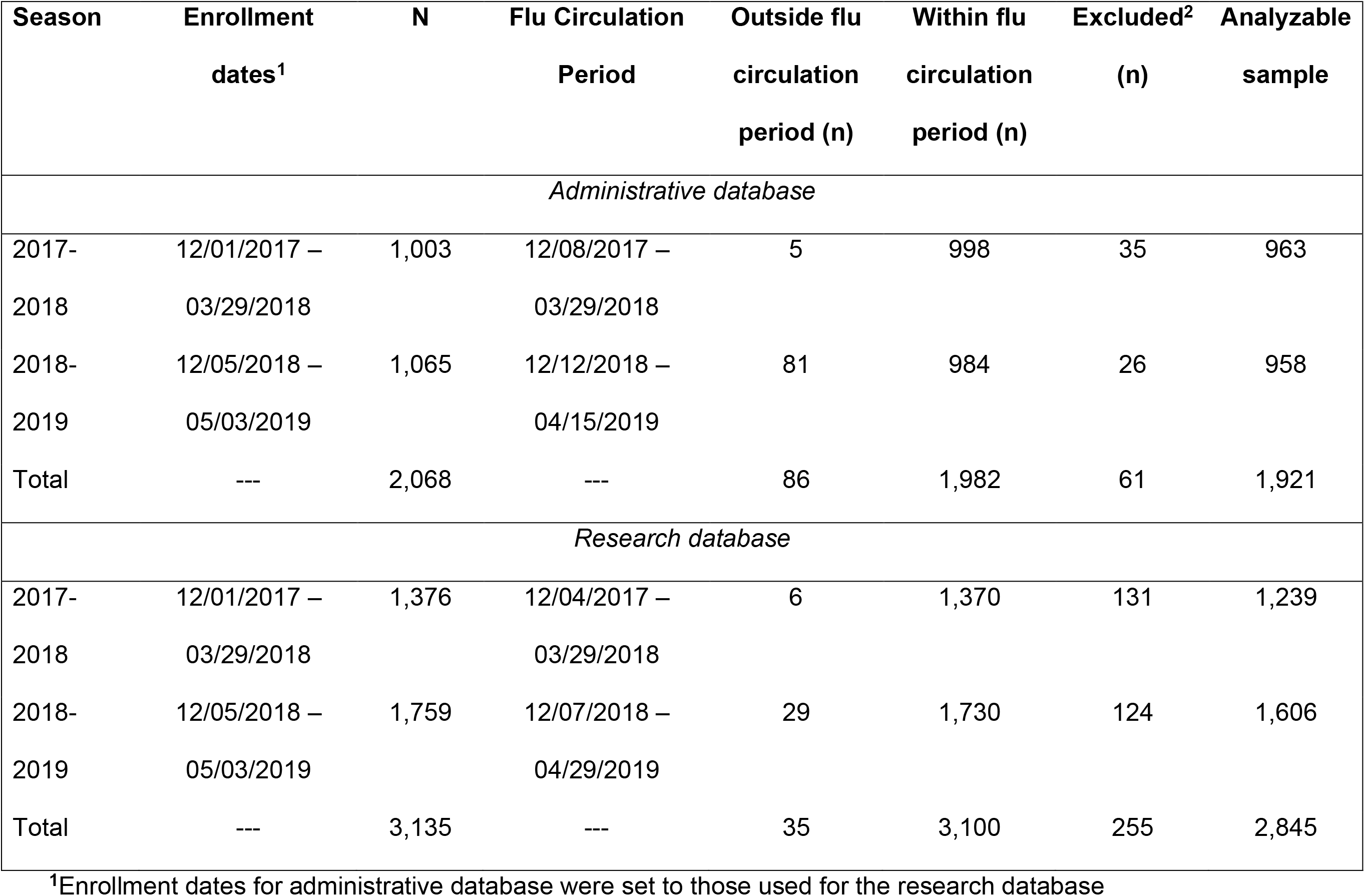
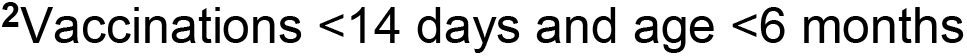
Description of analyzable sample from two sources.

| **Season** | **Enrollment**<br>**dates<sup>1</sup>** | **N** | **Flu Circulation**<br>**Period** | **Outside flu**<br>**circulation**<br>**period (n)** | **Within flu**<br>**circulation**<br>**period (n)** | **Excluded<sup>2</sup>**<br>**(n)** | **Analyzable**<br>**sample** |
| --- | --- | --- | --- | --- | --- | --- | --- |
| *Administrative database* |  |  |  |  |  |  |  |
| 2017-2018 | 12/01/2017 –<br>03/29/2018 | 1,003 | 12/08/2017 –<br>03/29/2018 | 5 | 998 | 35 | 963 |
| 2018-2019 | 12/05/2018 –<br>05/03/2019 | 1,065 | 12/12/2018 –<br>04/15/2019 | 81 | 984 | 26 | 958 |
| Total | --- | 2,068 | --- | 86 | 1,982 | 61 | 1,921 |
| *Research database* |  |  |  |  |  |  |  |
| 2017-2018 | 12/01/2017 –<br>03/29/2018 | 1,376 | 12/04/2017 –<br>03/29/2018 | 6 | 1,370 | 131 | 1,239 |
| 2018-2019 | 12/05/2018 –<br>05/03/2019 | 1,759 | 12/07/2018 –<br>04/29/2019 | 29 | 1,730 | 124 | 1,606 |
| Total | --- | 3,135 | --- | 35 | 3,100 | 255 | 2,845 |

<sup>1</sup>Enrollment dates for administrative database were set to those used for the research database
<sup>2</sup>Vaccinations <14 days and age <6 months

### Statistical analyses

Summary statistics of the demographic and clinical characteristics were determined for the administrative and research databases. Baseline characteristics were compared between vaccinated and unvaccinated patients using Chi-square tests for categorical variables.

A test-negative study estimates VE by comparing the odds of vaccination among RT-PCR confirmed influenza cases to the odds of vaccination among controls. Using odds ratios obtained from multivariable logistic regression models, VE estimates were calculated as (1-aOR) × 100.

A series of logistic regression models was conducted with RT-PCR confirmed influenza A and B as the dependent variables and vaccination status as the independent variable. The primary analyses determined VE for all influenza; subgroup analyses determined VE for influenza A/H1N1, influenza A/H3N2, and influenza B (both lineages combined due to small numbers of cases). The logistic regression models were adjusted *a priori* for age group (6 months-17 years, 18-49 years, 50-64 years and 65+ years), sex, race (white, non-white), influenza season (2017-2018, 2018-2019), prior vaccination status for the immediately preceding year and whether the visit took place in the emergency department. The VE and its 95% CI reported for the two databases were also stratified by age group and by season. Thus, age group was not adjusted for in the age-stratified model and season was not adjusted for in models which stratified seasons.

The significance of the difference between administrative and research database VE was identified through the effect of interactions in the logistic regression model. An indicator variable was created for database and the interaction of this binary indicator and the vaccination status was included in the model. Data were analyzed using SAS version 9.4 (SAS Institute, Cary, NC, USA). Statistical significance was defined as a two-sided *P* value <0.05. The University of Pittsburgh IRB approved the study.

## Results

There were significant differences in demographic characteristics between the administrative and research databases as shown in Table 2. For example, compared with those in the administrative database, research participants were more likely to be younger, vaccinated, previously vaccinated, infected with influenza A/H1N1 or A/H3N2, white and less likely to have been seen in the emergency department. Thus, the first difference between the methodologies is reflected in the demographics of the two populations.

**Table 2.** Characteristics of persons in administrative and research databases 2017-2019.

| Characteristic | Administrative database | Research database | *P* value | Administrative database vaccination status |  |  | Research database vaccination status |  |  |
| --- | --- | --- | --- | --- | --- | --- | --- | --- | --- |
|  | N=1,910 | N=2,845 |  | No<br>n=1,115 | Yes<br>n=795 | *P*<br>value | No<br>n=1,666 | Yes<br>n=1,179 | *P*<br>value |
| Age Group |  |  | **<0.001** |  |  | **<.001** |  |  | **<0.001** |
| 6 months – 17 years | 599 (31.4) | 986 (34.7) |  | 333 (29.9) | 266 (33.5) |  | 589 (35.3) | 397 (33.7) |  |
| 18 – 49 years | 677 (35.4) | 1,114 (39.1) |  | 457 (41.0) | 220 (27.7) |  | 756 (45.4) | 358 (30.4) |  |
| 50 - 64 years | 303 (15.9) | 451 (15.9) |  | 169 (15.2) | 134 (16.8) |  | 226 (13.6) | 225 (19.1) |  |
| ≥65 years | 331 (17.3) | 294 (10.3) |  | 156 (13.9) | 175 (22.0) |  | 95 (5.7) | 199 (16.8) |  |
| White race, ref. = non-white | 1,120 (58.6) | 1,958 (69.3) | **<0.001** | 573 (51.4) | 547 (68.8) | **<.001** | 1,080 (65.4) | 878 (74.7) | **<0.001** |
| Female sex, ref. = male | 1,108 (58.0) | 1,601 (56.3) | 0.241 | 649 (58.2) | 459 (57.7) | 0.837 | 881 (52.9) | 720 (61.1) | **<0.001** |
| Prior vaccination, ref. = not vaccinated prior year | 728 (38.1) | 1,123 (39.5) | 0.347 | 215 (19.3) | 513 (64.5) | **<.001** | 313 (18.8) | 810 (68.7) | **<0.001** |
| Emergency department testing/enrollment | 1,387 (72.1) | 987 (34.7) | **<0.001** | 857 (76.9) | 521 (65.5) | **<.001** | 621 (37.3) | 366 (31.0) | **<0.001** |
| Vaccinated, ref. = unvaccinated | 795 (41.6) | 1,179 (41.4) | 0.901 | -- | -- | -- | -- | -- | -- |

Table 2 also compares vaccinated with and non-vaccinated subjects within each database. In the administrative database, the vaccinated compared with the unvaccinated differed by age, race, (69% vs. 51% white; *P*<0.001), previous vaccination status (65% vs. 19%; *P*<0.001) and number of emergency department enrollments (66% vs. 77%; *P*<0.001). Among the individuals in the research database, compared with the unvaccinated, the vaccinated were older, more often white than non-white, female, previously vaccinated, and less frequently enrolled through the emergency department (all *P*<0.001).

Influenza circulation differed in the two seasons; influenza A/H3N2 was the predominant strain in 2017-2018, although there was circulation of A/H1N1 and influenza B, whereas in 2018-2019, A/H3N2 and A/H1N1 circulated in nearly equal proportions with a small proportion of influenza B. Table 3 shows the unadjusted and adjusted VE estimates from each data base for any influenza, influenza A, A/H1N1, A/H3N2, and influenza B for 2017-2018, 2018-2019, and for both seasons combined. Using the administrative database significant VE estimates were observed for any influenza, and influenza A for both seasons individually and combined, and for both influenza A/H1N1 and A/H3N2 for 2018-2018 and both seasons combined. VE for influenza B was not significant for either season singly or combined. Using the research database, VE was significant for all strains and substrains measured in each season and overall, with the exception of influenza B in 2018-2019. Significant VE estimates ranged from 39% (95%CI=15, 57) for A/H3N2 in 2017-2018 to 69% (95% CI=35, 85) for A/H1N1 for the 2017-2018 season. The last column in Table 3 indicates the *P* value for the comparison of VE estimates between both data sources. The research VE estimates were not significantly different from administrative VE estimates with the exception of influenza B in 2017-2018. Although not statistically significant due to overlapping confidence intervals and limited sample size, the VE for any influenza for 2017-18 was 12 percentage points higher for the research database (49%; 95%CI=31,62) than for the administrative database (37%; 95%CI=13, 54). The adjusted VE estimates for influenza B differed by >40 percentage points in 2017-2018 and the 2017-2019 combined seasons between the administrative and research VE estimates, but differences were not statistically significant.

**Table 3.**
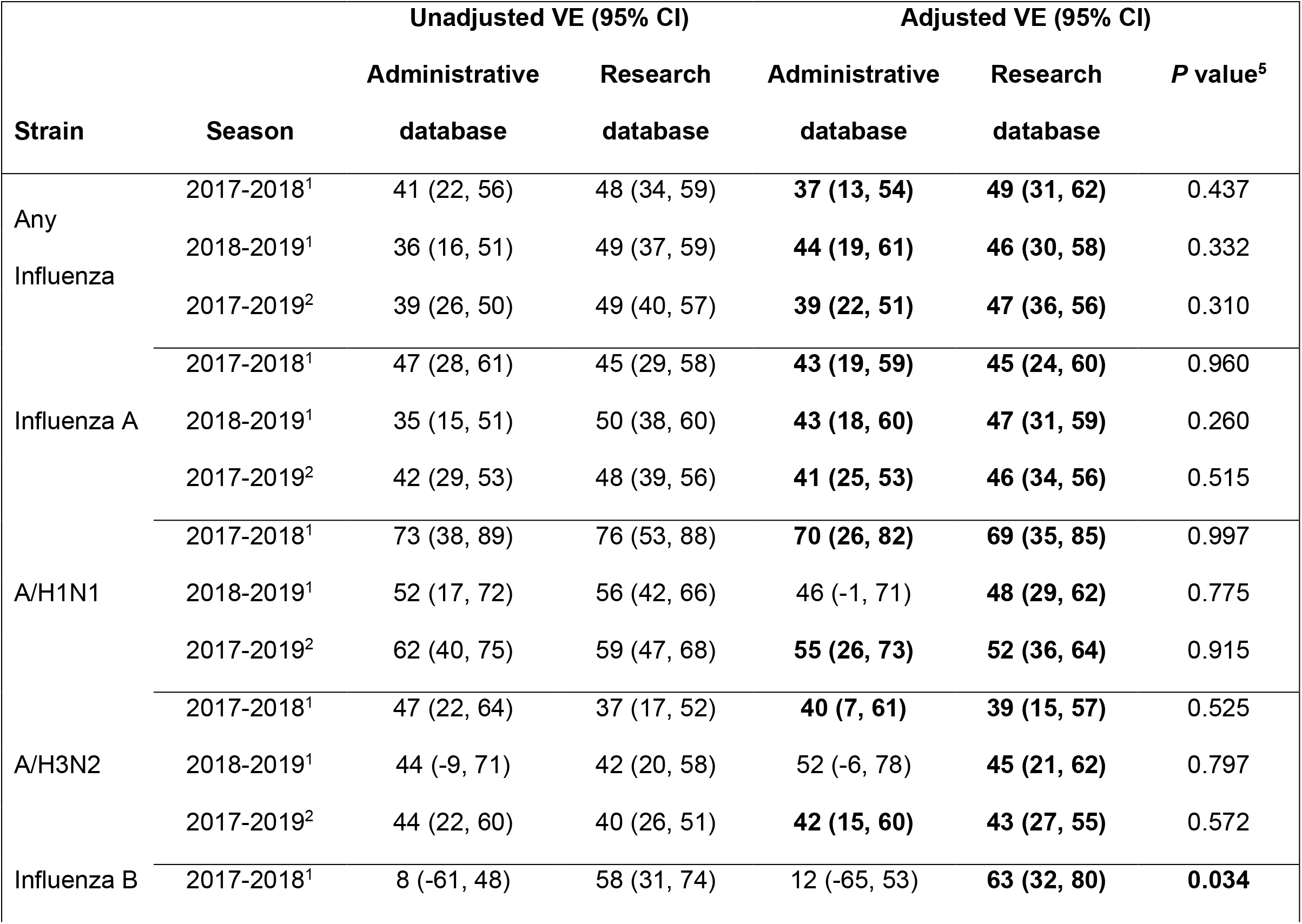
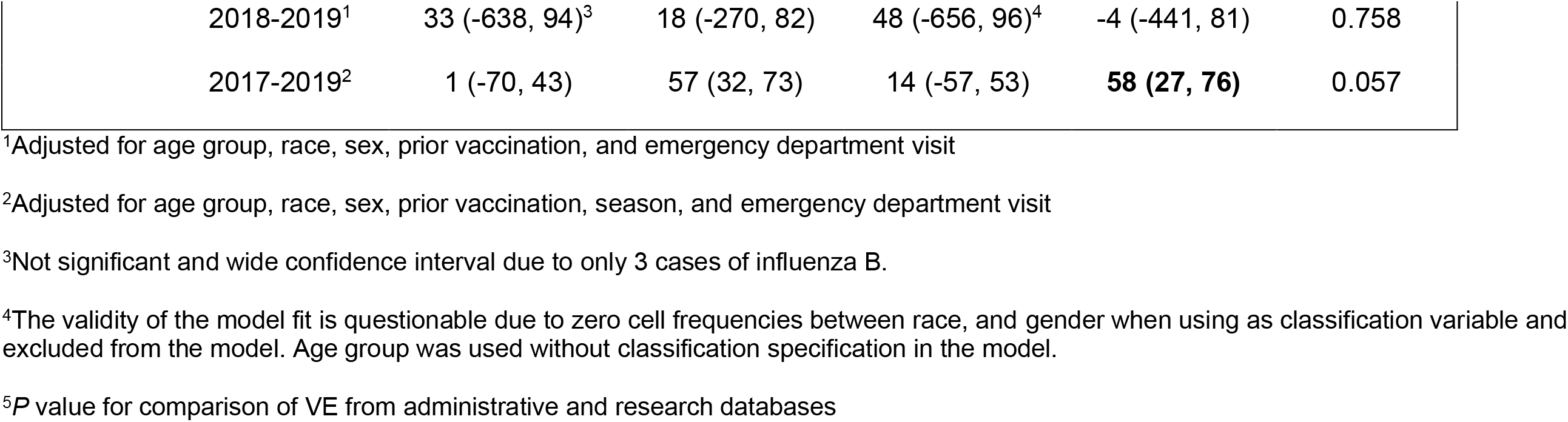
Comparison of vaccine effectiveness across all age-groups for 2017-2019 derived from administrative and research databases.

Data from both 2017-2018 and 2018-2019 were combined for VE analyses by age group shown in Supplemental Table 1. Significant VE estimates were observed consistently in the youngest age group (6 months-17 years) for any influenza (55% and 64%), influenza A (60% and 64%), influenza A/H1N1 (78% and 72%) and influenza A/H3N2 (78% and 56%). VE point estimates for 18-49-year-olds in the research database were significant for any influenza (41%), influenza A (35%), and A/H3N2 (36%).

Table 4 shows the demographic characteristics and VE estimates of patients in each of the databases split into emergency departments and outpatient clinics. In the emergency departments, patients in the administrative databases were significantly older (*P*<0.001), more often white (50.5% vs 43.9%; *P*=0.002) and more often female (58.5% vs. 49.1%; *P*<0.001) than those in the research database. Furthermore, when interaction terms were used, VE estimates from the administrative database were significantly lower than from the research database against all influenza (46%, 95% CI=29%, 59% vs. 66%, 95% CI=52%, 76%; *P*<0.001) and any influenza A (49%, 95% CI=33%, 62% vs. 67%, 95% CI=53,% 77%; *P*=0.002).

**Table 4.**
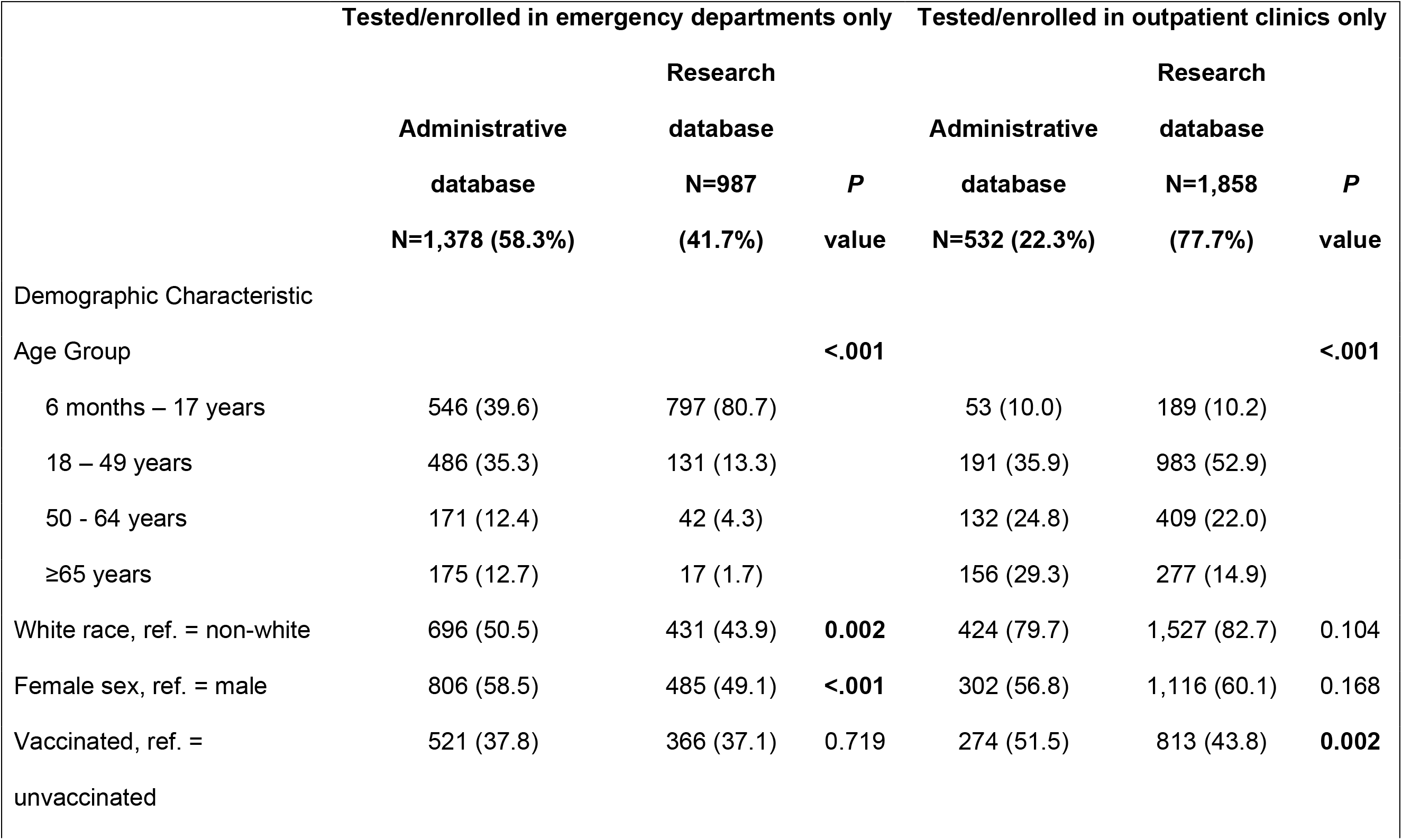
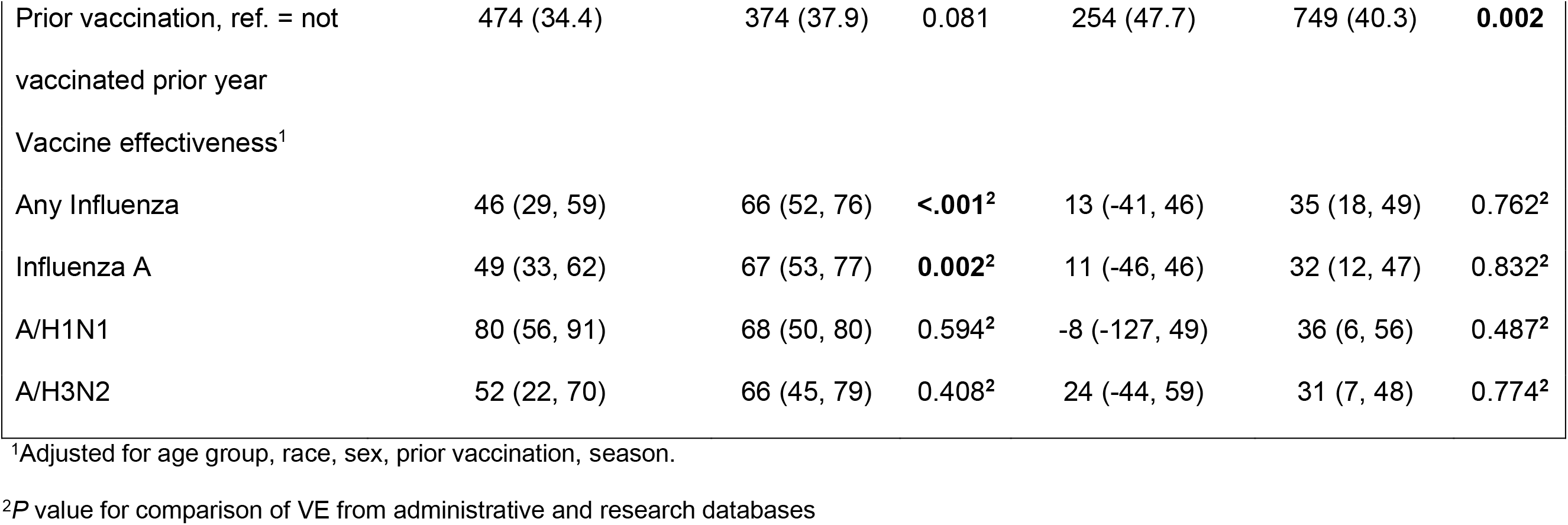
Characteristics and vaccine effectiveness estimates of persons in administrative and research databases 2017-2019.

Three differences between the databases for those enrolled in outpatient clinics were noted. Administrative database enrollees were significantly older (*P*<0.001), and more often vaccinated both in the enrollment season (51.5% vs. 43.8%; *P*=0.002) and in the prior season (47.7% vs. 40.3%; *P*=0.002).

## Discussion

In this comparison of influenza vaccine effectiveness estimates from a test-negative case control research study and administrative data sources, subjects differed by demographic characteristics of age, race and site of enrollment. The administrative database included older patients, more non-whites and more patients enrolled in the emergency department. With this limited sample size, the majority of comparisons found no significant differences in VE estimates between the two databases; however, differences were found for influenza B in 2017-2018 and with a larger sample size may have been found for others, given the differences in percentage points between estimates. When the databases were further divided into emergency department and outpatient clinics, new patterns emerged. In emergency departments, research enrollees were younger, more often male and non-white and VE against any influenza and all influenza A was significantly higher than was found in the administrative database. Whereas, in the outpatient clinics, age distribution was more evenly distributed in the administrative database and vaccination rates appeared to be higher than in the research database, but there were no significant differences in VE estimates.

The results of this study and examination of some of the literature suggest advantages and disadvantages of each type of data for determining influenza VE as shown in the box. Administrative database analysis for influenza vaccine effectiveness has some clear advantages that lie primarily in the potential sample sizes^4,10,11,12^ and relative cost per patient. Administrative databases can include large single health system data^2^ or can combine data across large geographic areas,^1^ thus may more accurately estimate VE for a country or region. These large datasets would be expected to be sufficiently powered to produce have narrow confidence intervals and thus instill confidence in their VE estimates. Large sample sizes, with rich EMR data, would also allow for inclusion of instrumental variables in the analysis.^10^ The per patient cost of acquiring data on large numbers of patients would be lower because the cost of influenza testing would be part of clinical care and would not be included as part of the study. Furthermore, there would be no need to hire research assistants to screen, consent and enroll participants. Administrative data collection does not carry risk of infection to the research staff that in-person enrollment does. Finally, the results of clinical influenza testing are likely to be rapidly available because of their importance to clinical care and infection control.

Conversely, use of administrative databases for VE analyses has some disadvantages. For example, administrative databases may represent the subgroup of individuals who seek care at the specific types of facilities contained in the database, such as the hospital-based clinics in this study. This situation may result in demographic, health or healthcare-seeking behavior characteristics and may limit general applicability of VE estimates. There may be limited information available about patients who are included without consent;^4^ the quality of the data received may vary; there may be delays in completing administrative databases,^4^ especially if data are being combined from several health systems; or there may not be indicators for factors that may affect VE in some groups, such as frailty among older adults.^13^ Selection biases are a major concern in administrative data-based, case-control studies for which making adequate adjustments can be difficult, leading to residual confounding.^17^ One study found that administrative data sets were “efficient” but yielded “highly questionable” estimates; in fact, adjustment by propensity score matching “exacerbated bias.”^11^

Regarding influenza testing, there may be several types of tests used with varying sensitivity and specificity, influenza subtyping may not be available, and there may be policies that favor or disfavor influenza testing in certain settings that may introduce bias.^14^ A study of inpatient testing in our locale did not find any association between influenza vaccination and clinical testing. However, clinical testing was significantly higher during the peak and post peak influenza periods than earlier in the season and higher among younger hospitalized patients with an ARI.^15^

In administrative databases, vaccine verification is limited to electronic sources. It is not known how many health systems are linked electronically to their state’s immunization registries, what the lag time is for making those data transfers or the completeness of the registries’ data. Lack of an automatic feed to the EMR, or incomplete registry data could introduce bias by vaccination status misclassification.^16^

Using research databases to determine influenza VE also has advantages and disadvantages. The primary advantage of research databases is control. For example, the researcher can determine the type or types of influenza testing to be used based on test characteristics, the clinical setting for recruitment, whether or not to oversample certain subpopulations. The researcher can set standards for data quality and ensure that those standards are met with training and monitoring of research staff and monitoring data completeness and quality as they are being collected. The data may be more informative and complete because subtyping of influenza virus can be conducted, and manual, as well as electronic, vaccine verification can be employed. Because misclassification of vaccination status can produce substantial biases in VE estimates, manual verification may be worth the extra effort and expense.

The primary disadvantages of research databases are cost and scale. Significant human and other resources are needed to train research staff; identify, approach, and enroll participants while risking infection; transport research specimens; and analyze them. These specimens may be batch analyzed, delaying return of results. Distance of sites from the research offices or the testing labs may limit geographical reach of the study and the number of sites may be limited by costs.

#### Box. Comparison of Administrative and Research Database Analyses

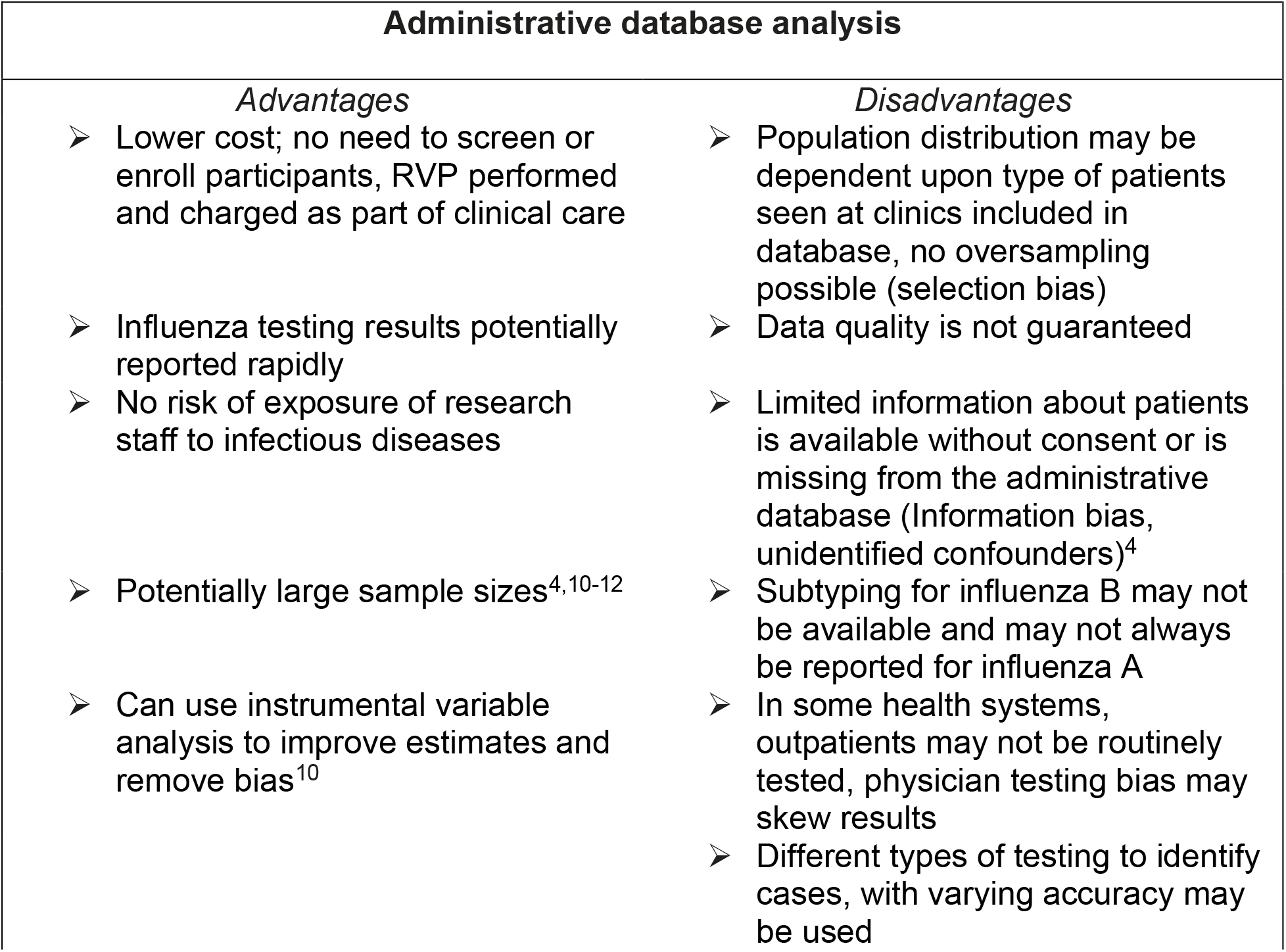
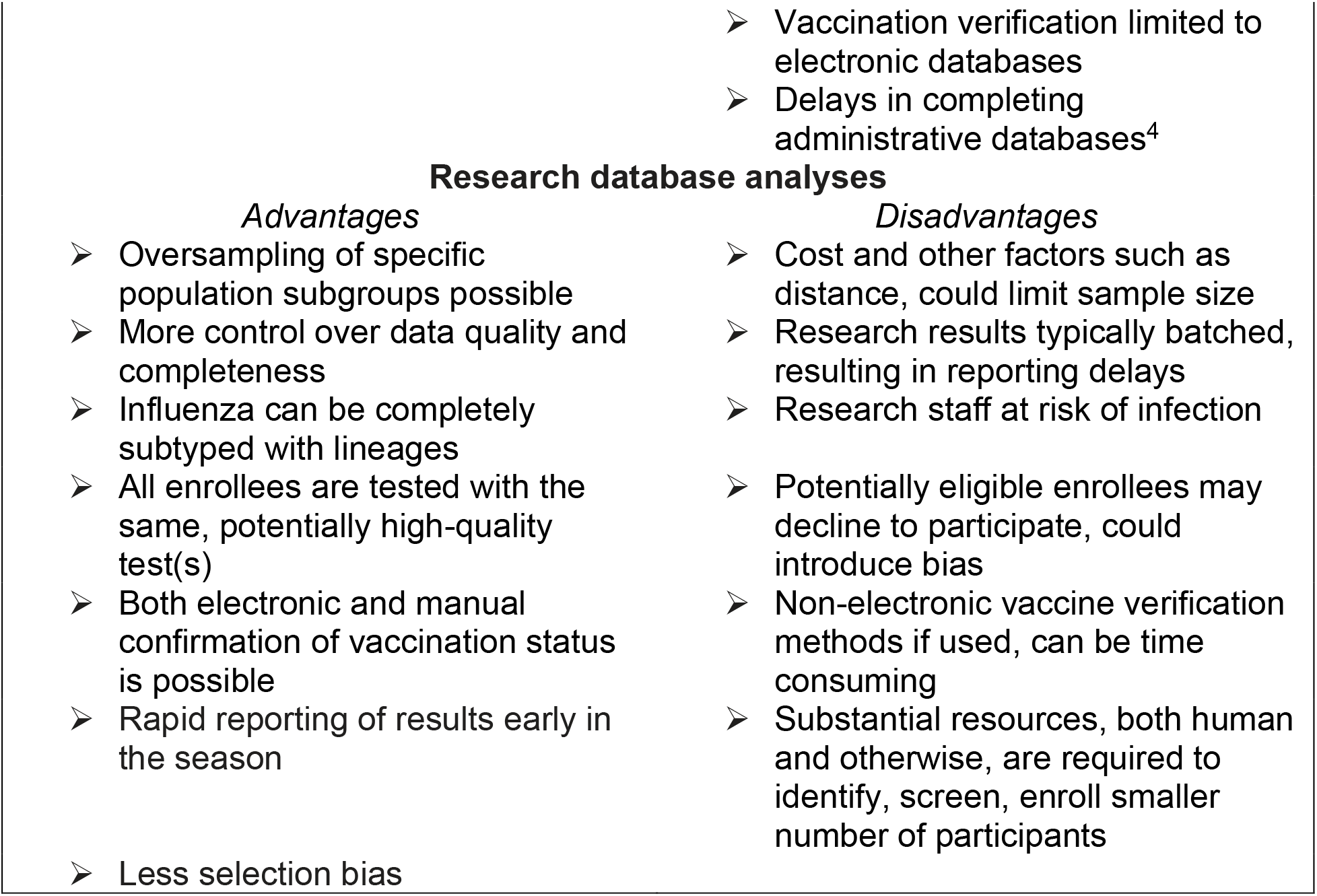

| **Administrative database analysis** |  |
| --- | --- |
| *Advantages* | *Disadvantages* |
| ➤ Lower cost; no need to screen or enroll participants, RVP performed and charged as part of clinical care<br>➤ Influenza testing results potentially reported rapidly<br>➤ No risk of exposure of research staff to infectious diseases<br>➤ Potentially large sample sizes<sup>4,10-12</sup><br>➤ Can use instrumental variable analysis to improve estimates and remove bias<sup>10</sup> | ➤ Population distribution may be dependent upon type of patients seen at clinics included in database, no oversampling possible (selection bias)<br>➤ Data quality is not guaranteed<br>➤ Limited information about patients is available without consent or is missing from the administrative database (Information bias, unidentified confounders)<sup>4</sup><br>➤ Subtyping for influenza B may not be available and may not always be reported for influenza A<br>➤ In some health systems, outpatients may not be routinely tested, physician testing bias may skew results<br>➤ Different types of testing to identify cases, with varying accuracy may be used |

No matter which method is used, speed to release of VE estimates depends on the relative timeliness of all parts of the process from data collection to analyses to compiling the report. If administrative data were readily available, they might be the best source for rapid influenza VE estimates, with the caveat that their generalizability may be limited. Currently the US Flu VE Network provides mid-season VE estimates using self-reported vaccination status that are published in the *Morbidity and Mortality Weekly Report*. Earlier VE estimates based on administrative databases may be useful in a severe influenza season, or in the present era of potential co-circulation of SARS-CoV-2, when treatment and infection control measures might differ.

### Strengths and Limitations

Strengths of this study include the test-negative design that utilizes the highly specific molecular methods to diagnose influenza, limiting misclassification of the outcome status. The sample size was limited, particularly for substrain analyses, as we found that point estimate differences of 40 were not significant, due to overlapping confidence intervals. Thus, the study was underpowered. The number of potential confounding factors included in regression modeling were limited due to the limited data available using administrative testing and vaccination databases. Whether residual confounding would differ between the administrative and research databases is unknown. Without additional sources of vaccination data, the differences in vaccination coverage between databases may be attributed to underreporting to the state registry, especially among adults. Incomplete vaccination data could result in inaccurate VE estimates. Finally, this study was conducted in one locale and should be repeated in other networks.

### Conclusions

The selection of the appropriate method for determining influenza vaccine effectiveness depends on a multitude of factors. Among those factors is sample size, which likely obscured significant differences between administrative and research database VE estimates in this study except when limiting the analyses to the emergency departments, in which case the research estimates were better. Differences in the types of persons enrolled in the databases, suggest that research estimates may be more generalizable. Other advantages of research databases for VE estimates include lack of clinician-related selection bias for testing and less misclassification of vaccination status because multiple sources are used. The advantages of the administrative databases are potentially shorter time to VE results and lower cost.

## Data Availability

Data may be made available after peer-reviewed publication.

---

**Supplemental Table 1.** Comparison of vaccine effectiveness in Pittsburgh for seasons 2017-2019 by age group, derived from administrative and research databases.

| Strain | Age group<br>(years) | Unadjusted VE (95% CI) |  | Adjusted VE (95% CI) |  |
| --- | --- | --- | --- | --- | --- |
|  |  | Administrative<br>database | Research<br>database | Administrative<br>database | Research<br>database |
| Any Influenza | 6 m- 17 | 53 (35, 66) | 65 (53, 74) | **55 (32, 70)** | **64 (50, 74)** |
|  | 18-49 | 40 (14, 58) | 46 (29, 59) | 32 (-2, 55) | **41 (19, 57)** |
|  | 50-64 | 18 (-39, 52) | 20 (-19, 46) | 17 (-51, 55) | 29 (-17, 57) |
|  | 65+ | 31 (-18, 59) | 23 (-30, 54) | 40 (-13, 68) | 7 (-77, 51) |
| Influenza A | 6 m- 17 | 56 (39, 69) | 66 (53, 75) | **60 (38, 74)** | **64 (50, 74)** |
|  | 18-49 | 40 (12, 58) | 41 (22, 55) | 32 (-3, 55) | **35 (10, 53)** |
|  | 50-64 | 20 (-40, 54) | 19 (-23, 47) | 22 (-47, 59) | 24 (-29, 55) |
|  | 65+ | 34 (-15, 62) | 25 (-31, 57) | 42 (-12, 70) | 14 (-67, 56) |
| A/H1N1 | 6 m- 17 | 80 (30, 95) | 76 (63, 84) | **78 (11, 94)** | **72 (56, 82)** |
|  | 18-49 | 38 (-16, 66) | 47 (18, 66) | 39 (-23, 70) | 36 (-6, 61) |
|  | 50-64 | 68 (12, 89) | 33 (-24, 64) | **70 (10, 90)** | 2 (-110, 54) |
|  | 65+ | 59 (-40, 88) | 28 (-61, 68) | 59 (-74, 90) | 11 (-160, 70) |
| A/H3N2 | 6 m- 17 | 81 (48, 93) | 53 (31, 68) | **78 (36, 93)** | **56 (33, 71)** |
|  | 18-49 | 34 (-13, 61) | 38 (14, 56) | 26 (-35, 59) | **36 (5, 57)** |
|  | 50-64 | 25 (-73, 67) | 11 (-47, 47) | 22 (-98, 70) | 38 (-20, 68) |
|  | 65+ | 33 (-24, 64) | 22 (-51, 60) | 43 (-18, 72) | 15 (-91, 62) |
| Influenza B | 6 m- 17 | -8 (-147, 52) | 67 (8, 88) | -7 (-166, 57) | 59 (-24, 87) |
|  | 18-49 | 51 (-77, 87) | 82 (49, 94) | 49 (-113, 88) | **81 (41, 94)<sup>3</sup>** |
|  | 50-64 | 4 (-267, 75) | 23 (-90, 69) | -34 (-484, 69) | 68 (-19, 91)<sup>3</sup> |
|  | 65+ | -10 (-401, 76) | 12 (-203, 74) | 3 (-484, 84) | -66 (-709, 66)<sup>3</sup> |

<sup>1</sup>P value for comparison of VE from administrative and research databases
<sup>2</sup> Adjusted for race, sex, season, prior vaccination, and emergency department visit. Season was excluded from the adjusted model. No influenza B cases in season 2018 for these age groups
<sup>3</sup>Emergency department visit data not available and excluded from the adjusted model

**Figure 1a.**
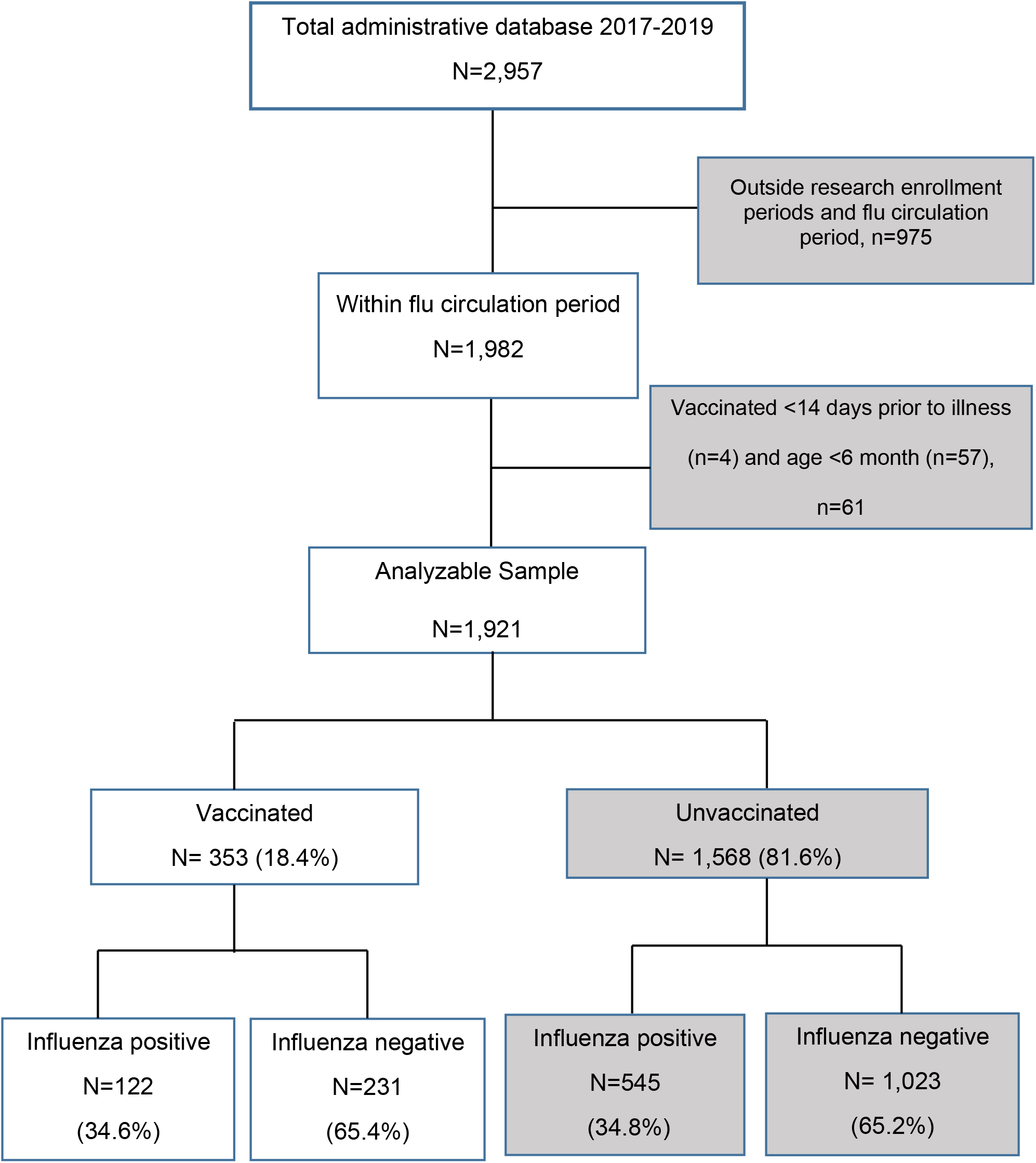
Flow chart for administrative database.

**Figure 1b.**
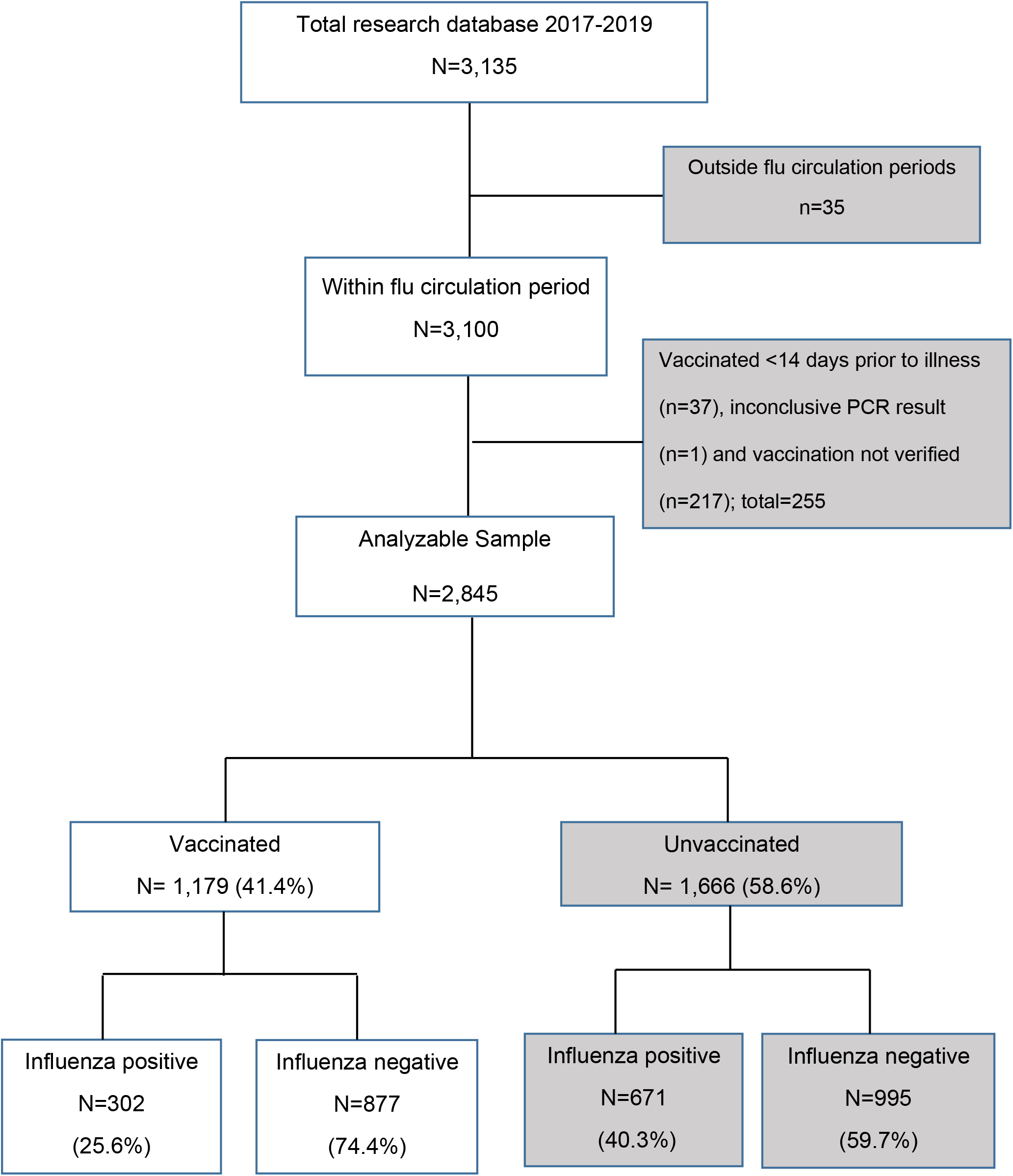
Flow chart for research data base.

## References

1. Zhou H, Thompson WW, Belongia EA, et al. Estimated rates of influenza-associated outpatient visits during 2001-2010 in 6 US integrated healthcare delivery organizations. Influenza and other respiratory viruses. 2018;12(1):122–131.

2. Temte JL, Barlow S, Schemmel A, et al. New method for real time influenza surveillance in primary care: a Wisconsin Research and Education Network (WREN) supported study. The Journal of the American Board of Family Medicine. 2017;30(5):615–623.

3. Thompson MG, Kwong JC, Regan AK, et al. Influenza vaccine effectiveness in preventing influenza-associated hospitalizations during pregnancy: a multi-country retrospective test negative design study, 2010-2016. Clinical Infectious Diseases. 2019;68(9):1444–1453.

4. Kwong JC, Buchan SA, Chung H, et al. Can routinely collected laboratory and health administrative data be used to assess influenza vaccine effectiveness? assessing the validity of the flu and other respiratory viruses research (forever) cohort. Vaccine. 2019;37(31):4392–4400.

5. Ohmit SE, Thompson MG, Petrie JG, et al. Influenza vaccine effectiveness in the 2011-2012 season: protection against each circulating virus and the effect of prior vaccination on estimates. Clinical infectious diseases: an official publication of the Infectious Diseases Society of America. 2014;58(3):319–327.

6. McLean HQ, Thompson MG, Sundaram ME, et al. Influenza vaccine effectiveness in the United States during 2012-2013: variable protection by age and virus type. The Journal of infectious diseases. 2015;211(10):1529–1540.

7. Gaglani M, Pruszynski J, Murthy K, et al. Influenza vaccine effectiveness against 2009 pandemic influenza A (H1N1) virus differed by vaccine type during 2013–2014 in the United States. Journal of Infectious Diseases. 2016;213(10):1546–1556.

8. Zimmerman RK, Nowalk MP, Chung J, et al. 2014–2015 influenza vaccine effectiveness in the United States by vaccine type. Clinical Infectious Diseases. 2016;63(12):1564–1573.

9. Jackson ML, Chung JR, Jackson LA, et al. Influenza Vaccine Effectiveness in the United States — 2015/16 Season. New England Journal of Medicine. 2017;377(6):534–543.

10. Wong K, Campitelli MA, Stukel TA, Kwong JC. Estimating influenza vaccine effectiveness in community-dwelling elderly patients using the instrumental variable analysis method. Archives of internal medicine. 2012;172(6):484–491.

11. Hottes TS, Skowronski DM, Hiebert B, et al. Influenza vaccine effectiveness in the elderly based on administrative databases: change in immunization habit as a marker for bias. PloS one. 2011;6(7):e22618.

12. Naleway AL, Ball S, Kwong JC, et al. Estimating vaccine effectiveness against hospitalized influenza during pregnancy: multicountry protocol for a retrospective cohort study. JMIR research protocols. 2019;8(1):e11333.

13. Jackson LA, Nelson JC, Benson P, et al. Functional status is a confounder of the association of influenza vaccine and risk of all cause mortality in seniors. Int J Epidemiol. 2006;35(2):345–352.

14. Skowronski DM, De Serres G, Orenstein WA. Caution required in the use of administrative data and general laboratory submissions for influenza vaccine effectiveness estimation. Clinical infectious diseases: an official publication of the Infectious Diseases Society of America. 2019;69(6):1084.

15. Balasubramani GK, Saul S, Nowalk MP, Middleton DB, Ferdinands JM, Zimmerman RK. Does influenza vaccination status change physician ordering patterns for respiratory viral panels? Inspection for selection bias. Human vaccines & immunotherapeutics. 2019;15(1):91–96.

16. Orenstein WA, Bernier RH, Dondero TJ, et al. Field evaluation of vaccine efficacy. Bulletin of the World Health Organization. 1985;63(6):1055.

17. Gavrielov-Yusim, N., & Friger, M. (2014). Use of administrative medical databases in population-based research. Journal of epidemiology and community health, 68(3), 283–287. https://doi.org/10.1136/jech-2013-202744

